# C-NODDI: a constrained NODDI model for axonal density and orientation determinations in cerebral white matter in normative aging

**DOI:** 10.1101/2023.03.06.23286818

**Authors:** Maryam H. Alsameen, Zhaoyuan Gong, Wenshu Qian, Matthew Kiely, Curtis Triebswetter, Christopher M. Bergeron, Luis E. Cortina, Mary E. Faulkner, John P. Laporte, Mustapha Bouhrara

## Abstract

**Purpose:** Neurite orientation dispersion and density imaging (NODDI) provides measures of neurite density and dispersion through computation of the neurite density index (NDI) and the orientation dispersion index (ODI). However, NODDI overestimates the cerebrospinal fluid water fraction in white matter (WM) and provides physiologically unrealistic high NDI values. Furthermore, derived NDI values are echo time (TE)-dependent. In this work, we propose a modification of NODDI, named constrained NODDI (C-NODDI), for NDI and ODI mapping in WM.

**Methods:** Using NODDI and C-NODDI, we investigated age-related alterations in WM in a cohort of 58 cognitively unimpaired adults. Further, NDI values derived using NODDI or C-NODDI were correlated with the neurofilament light chain (NfL) concentration levels, a plasma biomarker of axonal degeneration.

**Results:** ODI derived values using both approaches were virtually identical. We confirm the previous finding that NDI estimation using NODDI is TE-dependent. In contrast, C-NODDI-NDI exhibit lower susceptibility to TE. Further, C-NODDI-NDI values were lower than NODDI-NDI. Further, our results indicate a quadratic relationship between NDI and age suggesting that axonal maturation continues until middle age followed by a decrease. This quadratic association was notably significant in several WM regions using C-NODDI, while limited to a few regions using NODDI. ODI exhibited overall constant trends with age. Finally, C-NODDI-NDI values exhibited a stronger correlation with NfL concentration levels as compared NODDI-NDI, with lower NDI values correspond to higher levels of NfL.

**Conclusions:** C-NODDI provides a complementary method to NODDI for determination of NDI in white matter in normative aging.

## INTRODUCTION

Postmortem histological investigations have shown that cerebral tissue undergoes continuous microstructural and architectural changes throughout the lifespan (1–3). It has been suggested that axonal degeneration is among the main sequelae of aging as well as several age-related disorders, with concomitant motor and cognitive declines (4–11). Therefore, it is indispensable to characterize changes in axonal density that occur with normative aging to identify alterations arising from pathological manifestations. Whilst providing insights into cerebral gray matter (GM) and white matter (WM) maturation and degeneration, histological investigations cannot be performed in real-time on living subjects precluding longitudinal evaluations of brain aging, correlative studies with physical and cognitive performance, or interventions.

Magnetic resonance imaging (MRI), particularly diffusion tensor imaging (DTI), has been extensively used to investigate brain maturation and degeneration, suggesting complex and nonlinear trajectories of the DTI-based indices with age in WM and GM (12–18). Although DTI-indices are sensitive to the fraction of intracellular water, a proxy of axonal density, they are also sensitive to other tissue properties, such as fiber crossing and fanning, while lacking specificity to different diffusion tissue compartments. To overcome this difficulty, the neurite orientation dispersion and density imaging (NODDI) MRI technique has been introduced, providing measures of neurite density and dispersion through computation of the orientation dispersion index (ODI) and the neurite density index (NDI) (19). NODDI has been extensively used in clinical and preclinical studies of aging and neurological disorders (20–37). However, NODDI-based studies of axonal density and dispersion with normative aging remain limited, and with disparate results. Indeed, Billiet and colleagues and Chang and colleagues observed higher NDI values with age in several cerebral WM regions (38, 39), Merluzzi and colleagues’ observed lower NDI values in different cerebral WM structures (40), while Bouhrara and colleagues and Beck and colleagues have recently shown a complex regional association between NDI and age, with several cerebral structures exhibiting inverted U-shaped relationships. These observations suggest an increase in axonal density until middle age followed by a loss afterwards (6, 7). It remains unclear whether this discrepancy is due to differences in cohort characteristics or the experimental implementation of NODDI, including variations in echo-time (TE) (41).

NODDI is based on a multicompartmental model of water diffusion incorporating intracellular water, that is, water within neurites, extracellular water, and a compartment consisting of less restricted water from the cerebrospinal fluid (CSF) volume. Although NODDI has gained rapid popularity, criticisms were raised for its overestimation of the isotropically diffusing water fraction (*f*_iso_) of the CSF compartment and for providing unrealistically high NDI values in WM (19, 41, 42). Newly, Gong and colleagues have shown that derived NDI values from NODDI are dependent on the echo time (TE) (41). These drawbacks, hampering result interpretation and precluding multisite comparisons, are believed to be due to the underlying assumption in the original NODDI signal model where all compartments are considered to have similar transverse relaxation (*T_2_*) values (41, 42). Indeed, in recent works, Bouyagoub and colleagues have suggested rescaling *f*_iso_ using predetermined *T_2_* values of the CSF and intra/extracellular water compartments (42), while Gong and colleagues proposed a multi-echo time NODDI (MTE-NODDI) approach incorporating several NODDI scans performed at different TEs (41). Although these compelling advanced approaches have led to plausible NDI values in WM, they require a lengthy extension of the total scan time making them hardly practical in clinical setting.

In this work, we propose a modification of NODDI that requires no extension of the acquisition time. Our approach is based on the modification of the NODDI signal model such that *f*_iso_ is provided as an input (*i.e.,* constrained) value in each voxel. This bicomponent model simplifies the tricomponent model used in the original NODDI. We named this approach constrained NODDI (C-NODDI). Unlike Gong and colleagues’ MTE-NODDI model, C-NODDI assumes identical *T_2_* values for both the intra and extra-cellular waters, in line with Bouyagoub and colleagues’ original formulation (42). Indeed, this assumption is supported by extensive evidence from previous relaxometry studies, demonstrating that the relaxation times of these two compartments are virtually similar (43–46). Using the original NODDI and C-NODDI approaches, we investigated age and sex-related microstructural alterations in WM in a cohort of 58 cognitively unimpaired adults. We also investigated the impact of TE on derived NDI and ODI values using NODDI and C-NODDI, and compared the results to those derived using MTE-NODDI. Finally, we compared the correlations between NDI derived values using NODDI or C-NODDI with the neurofilament light chain (NfL) concentration levels, a plasma biomarker of axonal degeneration, obtained from a subset of 43 participants included in our study cohort.

## MATERIALS & METHODS

### Participants

Participants were recruited from the Baltimore Longitudinal Study of Aging (BLSA) and the Genetic and Epigenetic Signatures of Translational Aging Laboratory Testing (GESTALT) (47, 48). BLSA and GESTALT are longitudinal cohort studies funded and conducted by the National Institute on Aging Intramural Research Program to evaluate multiple biomarkers related to aging. The inclusion and exclusion criteria for these two studies are essentially identical. Participants underwent a battery of cognitive tests and those with cognitive impairment, metallic implants, neurologic, or significant medical disorders were excluded (49). The final cohort consisted of 58 cognitively unimpaired volunteers (mean ± standard deviation of Mini-Mental State Examination (MMSE) = 29.2 ± 1.0) ranging in age from 21 to 83 years (45.4 ± 18.3 years), including 31 men (42.9 ± 17.5 years) and 27 women (48.3 ± 19.1 years). Age and MMSE did not differ significantly between men and women. The protocol was approved by the MedStar Research Institute and the National Institutes of Health Intramural Ethics committees, and all examinations were performed in compliance with the standards established by the National Institutes of Health Institutional Review Board. All participants provided written informed consents.

### Data acquisition

All experiments were performed with a 3T whole body Philips MRI system (Achieva, Best, The Netherlands) using the internal quadrature body coil for transmission and an eight-channel phased-array head coil for reception. Diffusion-weighted images (DWI) were acquired using a single-shot EPI sequence with repetition time (TR) of 10 s, echo time (TE) of 67 ms, three *b*-values of 0, 700, and 2000 s/mm^2^, with the two later encoded in 32 diffusion-weighting gradient directions, field-of-view (FoV) of 240 mm × 208 mm × 150 mm, acquisition matrix of 120 × 120 × 50, and acquisition voxel size of 2 mm × 2 mm × 3 mm. Two images at *b*-value of 0 s/mm^2^ were acquired. Images were acquired with SENSE factor of 2 and reconstructed to a voxel size of 2 mm × 2 mm × 2 mm. The total acquisition time was ∼16 min.

Finally, to evaluate the effect of TE on NDI and ODI derived using NODDI or C-NODDI, DW images were acquired for three subjects. Subject 1&2: DW images were acquired at four different TEs of 78, 90, 100 or 120 ms. For each TE, delta/DELTA values were set to 28.2/38.5 ms for TE = 78 ms, 34.2/44.5 ms for TE = 90 ms, 39.2/49.5 ms for TE = 100 ms and 45.2/55.5 for TE = 120 ms. Here again, three *b*-values of 0, 700, and 2000 s/mm^2^ were acquired, with the two later encoded in 32 diffusion-weighting gradient directions sampling the same *q*-space, FoV of 240 mm × 208 mm × 150 mm, acquisition matrix of 120 × 120 × 50, and acquisition voxel size of 2 mm × 2 mm × 3 mm. Subject 3: DW images were acquired at seven different TEs of 77, 87, 97, 107, 117, 127 and 137 ms. For each TE. Four *b*-values of 0, 700, 2000 and 3000 s/mm^2^ were acquired, with the three later encoded in 32 diffusion-weighting gradient directions sampling the same *q*-space. To accommodate the prolonged scan time, the FoV was set to 240 mm × 204 mm × 111 mm, acquisition matrix of 80 × 68 × 19, acquisition voxel size of 3 mm × 3 mm × 3 mm, and slice gap of 3 mm.

### The C-NODDI signal model

#### Signal model

NODDI is a multicompartmental signal model of water diffusion incorporating intracellular water, extracellular water and CSF water volumes (19). Assuming that each of these three compartments exhibits unique transverse and longitudinal relaxation times and diffusion, the signal model can be expressed as:

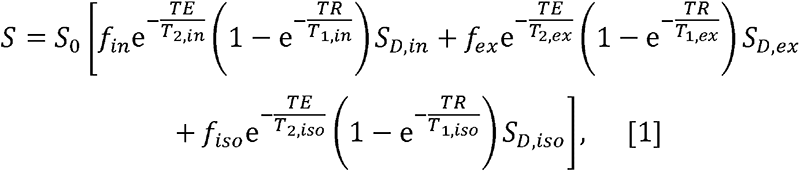

where *S* is the measured signal at a given combination of TE, repetition time (TR) and *b*-value; *S*_0_ is the signal at TE = 0 ms with TR = +∞ and *b*-value = 0 s/mm^2^; f_*in*_, f_*ex*_ and f_*iso*_ are, respectively, the fractions of the intracellular, extracellular and CSF water pools;T_*2, in*_, T_*2,ex*_ and T_*2,iso*_ are, respectively, the transverse relaxation times of the intracellular, extracellular and CSF water pools; T_*1,in*_, T_*1,ex*_ and T_*1,iso*_ are, respectively, the longitudinal relaxation times of the intracellular, extracellular and CSF water pools; and S_*D*_,in, S_*D,ex*_ and S_*d,iso*_, are, respectively, the signal attenuation due to water diffusion of the intracellular, extracellular and CSF water pools. When long TR is applied (*e.g.*, TR ≥ 5 s), the *T1* effect is mitigated so that Eq. 1 can be reduced to:

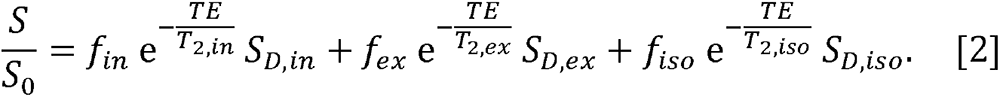

Furthermore, it has previously been shown that 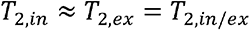 (43–46), in which case Eq. 2 can be reduced to:

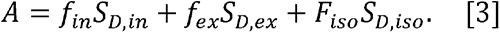

Where A=*S/S_b=0_* is the measured normalized diffusion-weighted signal, with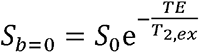 representing the *T_2_*-weigthed image obtained at *b*-value of 0 s/mm^2^, and 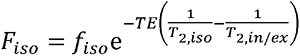. In voxels with no CSF contamination such as the deep WM regions,*f*_iso_= 0 so that Eq. 3 can be reduced to 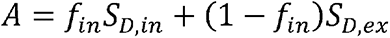, in which case, derived NDI (*i.e., f_in)_* values are expected to be relatively independent of *T2* and, therefore, are also relatively independent of the choice of TE. However, in regions with CSF contamination, *Fiso* > 0

#### NDI and ODI mapping

In the original NODDI approach, *F*_iso_ is derived along with NDI and ODI (19). We modified the NODDI MATLAB toolbox (https://www.nitrc.org/projects/noddi_toolbox) so that *F*_iso_ is instead provided as a known input parameter. This strategy allows a reduction of the number of unknown parameters to be estimated (Eq. 3), thus restricting the parameter space and improving the fitting stability of the C-NODDI signal model (50–53). Here, the *F*_iso_ map is computed from the *T_2_*-weigthed image obtained at *b*-value of 0 s/mm^2^ using the hidden Markov random field model and the expectation-maximization algorithm (54), known as FAST in the FSL software (55). FAST segments a structural image of the brain into different tissue classes, including WM, GM and CSF/*F*_iso_, providing, in each voxel, estimates of the fractions of these tissue compartments. Extensive work has previously been conducted to evaluate the accuracy of FAST for tissue segmentation (56–58). The derived *F*_iso_ map is then used as an input to calculate NDI and ODI using our modified NODDI approach, C-NODDI. All MATLAB codes are available upon request from the corresponding author.

For each participant, corresponding NDI and ODI maps were generated using NODDI and C-NODDI. Specifically, all DW images were registered to the *b_0_* image and corrected for motion and eddy current distortion artefacts using the Artefact Correction in Diffusion MRI (ACID) toolbox (http://diffusiontools.com/) (59). The co-registered DW images were then used to calculate NDI and ODI using NODDI or C-NODDI. NDI was defined as the *f_in_* for both approaches.

### Regions-of-interest determination

For each participant, using FSL, the DW image obtained with *b* of 0 s/mm^2^ was nonlinearly registered to the Montreal Neurological Institute (MNI) standard space and the derived transformation matrix was then applied to the corresponding NDI and ODI maps. Fourteen WM regions of interest (ROIs) were defined from MNI encompassing the whole brain (WB), frontal lobes (FL), parietal lobes (PL), temporal lobes (TL), occipital lobes (OL), cerebellum (CRB), corpus callosum (CC), internal capsules (IC), cerebral peduncle (CP), corona radiata (CR), thalamic radiation (TR), fronto-occipital fasciculus (FOF), longitudinal fasciculus (LF), and forceps (FR), as shown in Fig. 1. All ROIs were eroded to reduce partial volume effects and imperfect image registration using the FSL tool *fslmaths.* Finally, the mean NDI and ODI values within each ROI were calculated.

**Figure 1.**
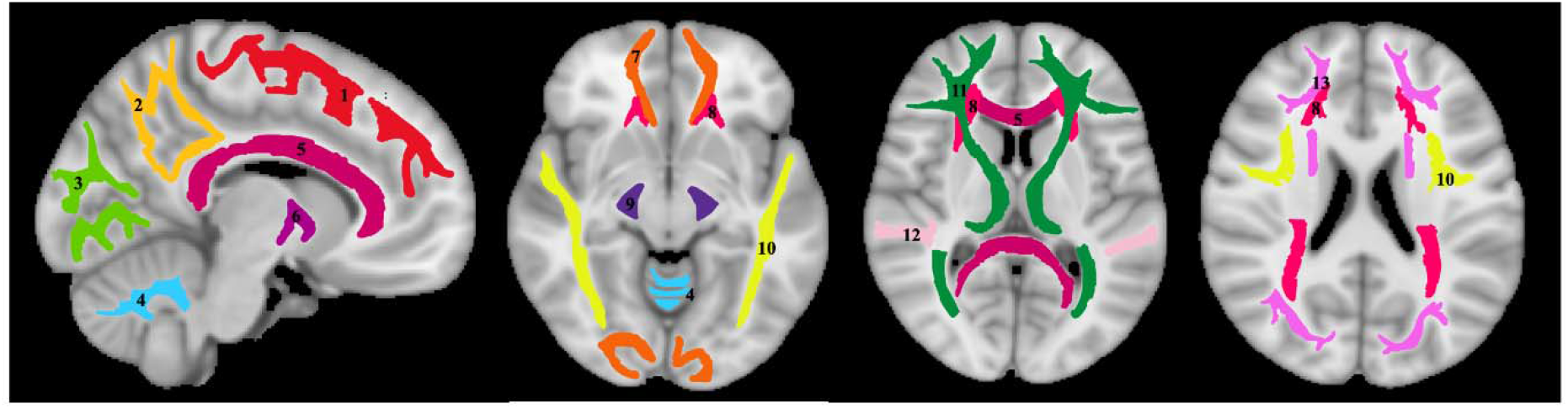
Visualization of the white matter ROIs used in our analysis. 1) Frontal lobes (FL), 2) parietal lobes (PL), 3) occipital lobes (OL), 4) cerebellum (CRB), 5) corpus callosum (CC), 6) internal capsules (IC), 7) forceps (FR), 8) corona radiata (CR), 9) cerebral peduncles (CP), 10) longitudinal fasciculus (LF), 11) thalamic radiation (TR), 12) temporal lobes (TL), 13) fronto-occipital fasciculus (FOF).

### Analyses

#### Differences in Fiso

in this analysis, we compared derived *F_iso_* maps using NODDI or FAST. Derived *F_iso_* maps were shown for a representative example.

#### Differences in NDI and ODI

in this analysis, we compared NDI and ODI maps derived using NODDI or C-NODDI. Average NDI and ODI maps by age intervals over the adult lifespan for a representative axial slice were calculated. Furthermore, to investigate the effects of age and sex on NDI and ODI, multiple linear regression analysis was applied using the mean NDI or ODI derived from NODDI or C-NODDI within each ROI as the dependent variable and sex, age, and age^2^ as the independent variables, after mean age centering. In all cases, the interactions between sex and age or age^2^ were found to be nonsignificant and were therefore omitted from the parsimonious model. Further, for each ROI, Pearson correlation analysis was conducted to examine the discrepancy between ODI, or NDI, values derived using NODDI or C-NODDI. For all analyses, the threshold for statistical significance was *p* < .05 after correction for multiple ROI comparisons using the FDR method (60, 61).

#### Correlations of NDI and NfL

in this analysis, we assessed the Pearson correlations between NDI derived using NODDI or C-NODDI and NfL which represents a plasma biomarker of axonal degeneration. NfL was measured from 43 participants of our study cohort (age: 50 [SD 18]). Blood for plasma biomarker measurement was collected at the time of MRI. Plasma was separated, aliquoted and stored at -80°C using standardized protocols. EDTA plasma was used to measure NfL using the Quanterix Single Molecule Array (Simoa) Neurology 4-Plex E assay on the HD-X Instrument (Quanterix Corporation).

#### Effect of TE

to investigate the effect of TE on derived NDI and ODI values using C-NODDI or NODDI, parameter maps as well as mean values calculated over a large WM region encompassing the cerebral lobes, at each TE, were displayed for three participants. Further, for each participant, we generated voxel-wise difference maps derived by subtracting each NDI, or ODI, map calculated at a given TE from the NDI, or ODI, map obtained at the lowest TE. Finally, the fitting quality was estimated in each voxel using the Bayesian information criterion (BIC) over all encoding directions for both NODDI and C-NODDI approaches at each TE, accounting for the differences in the complexity of these models (62). For each voxel, the ratio of these BCIs, given by Ratio-BIC = BIC-C-NODD / BIC-NODDI, was calculated.

#### NDI from MTE-NODDI

Using the MTE-NODDI analysis (41), for each participant, an NDI map was derived using the NODDI scan obtained at different TEs.

## RESULTS

Figure 2 shows representative *F*_iso_ maps derived using NODDI or FAST/FSL. It is readily seen that the *F*_iso_ map derived using NODDI exhibits abnormally high *F*_iso_ values, especially within several WM regions, with values reaching over 0.25 (*i.e.*, 25%) in several voxels. In contrast, derived *F*_iso_ values using the hidden Markov random field model and the expectation-maximization algorithm, the FAST algorithm as implemented in FSL, are within the physiologically expected ranges with values near zero in the deep WM regions.

**Figure 2:**
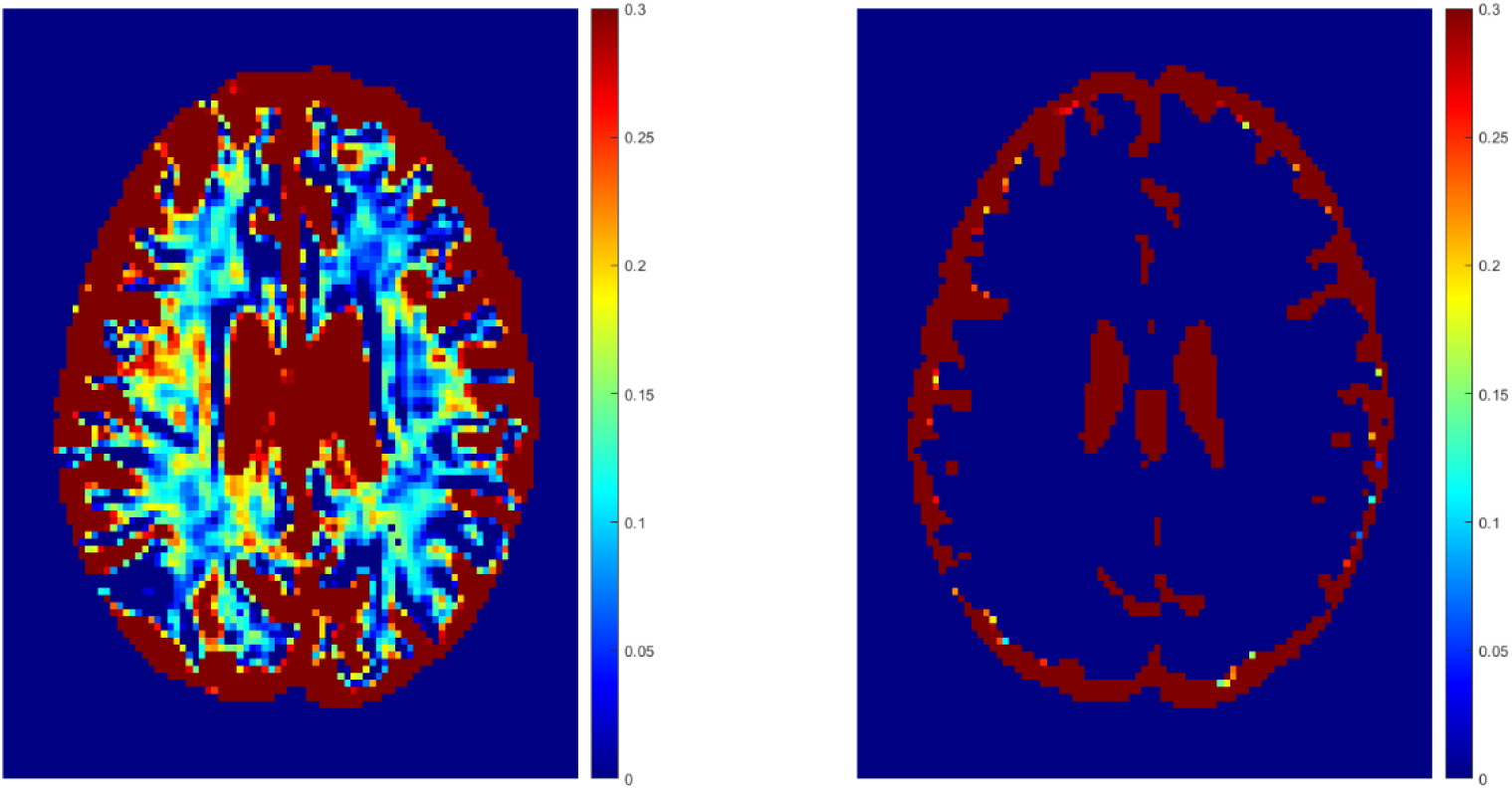
Representative maps of the fraction of the isotropic diffusion component (F_iso_), that is, the cerebrospinal fluid fraction, derived using NODDI (left image) or using the FAST algorithm as implemented in the FSL software (right image). It is readily seen that the NODDI approach overestimates F_iso_, especially in the white matter regions.

Figure 3a shows representative average NDI and ODI maps for different age intervals corresponding to young, middle, late middle and late adulthood. Visual inspection indicates increases in NDI values from early adulthood through middle age (*i.e.,* 40-59 years), followed by lower NDI values in several brain regions, consistent with progressive increases in axonal density followed by reductions at older ages. Furthermore, we note that different regions exhibit different patterns in the association of NDI with age. In contrast, the ODI maps exhibit low regional variations with age. Remarkably, NDI maps derived from NODDI exhibit values that exceed 0.7 (*i.e*., 70%) in several cerebral WM structures, while the NDI values derived using C-NODDI are considerably lower. Moreover, ODI maps derived using NODDI and C-NODDI were virtually identical. Indeed, our quantitative comparison, presented in Fig. 3b, indicates weak to moderate regional Pearson correlation between the NDI values derived using NODDI and those derived using C-NODDI for all ROIs investigated, in agreement with visual inspection (Fig. 3a). In contrast, strong regional correlations were observed between the ODI values derived from the two NODDI approaches.

**Figure 3a.**
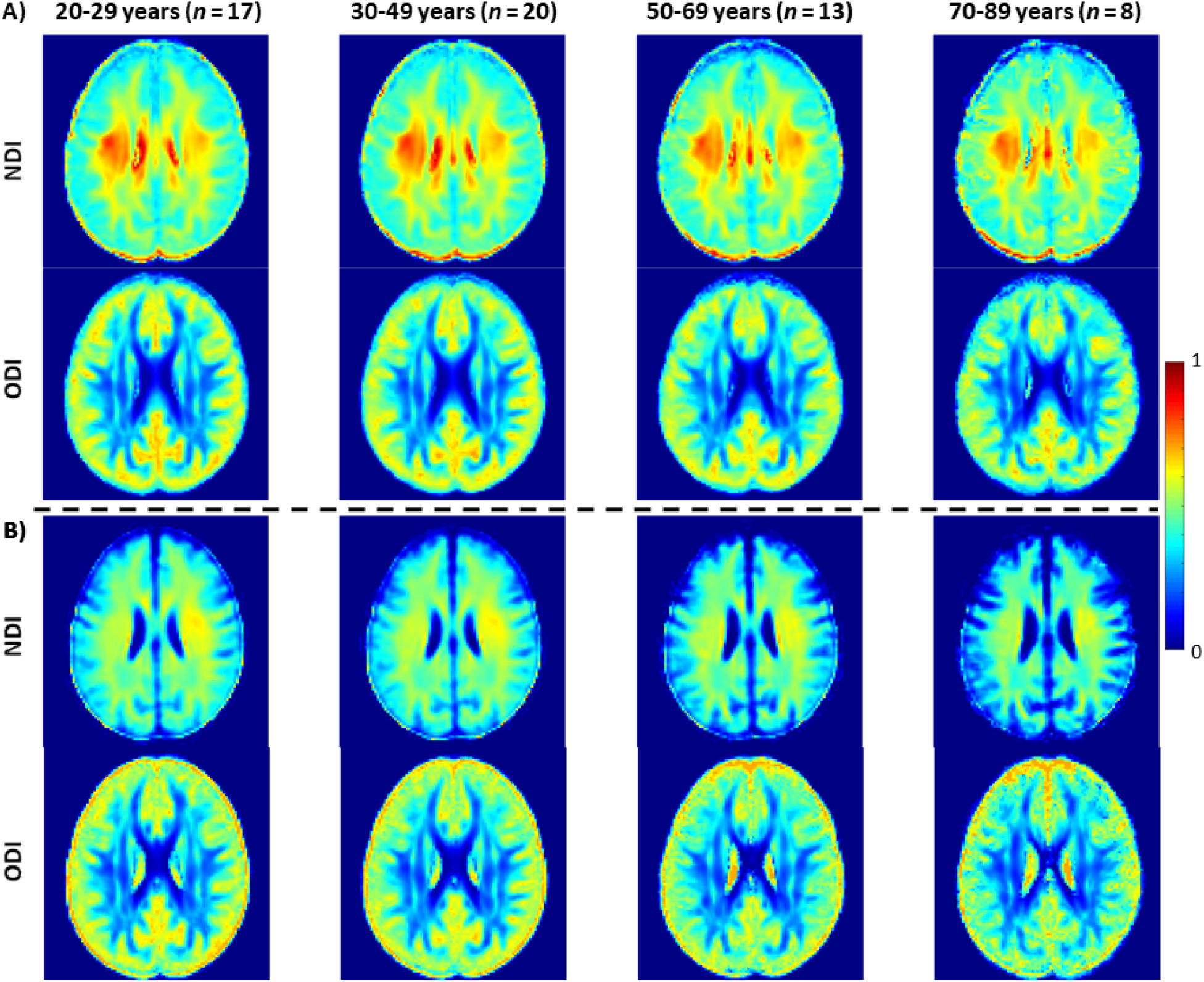
NDI and ODI maps were represented as averaged participant maps calculated over four age intervals. Results are shown for a representative slice. NDI and ODI maps derived A) using the original NODDI approach, and B) using the C-NODDI approach. While the ODI values derived using both approaches are virtually identical, C-NODDI provides substantially lower NDI values than those derived using NODDI, resulting in fewer unrealistic values.

**Figure 3b.**
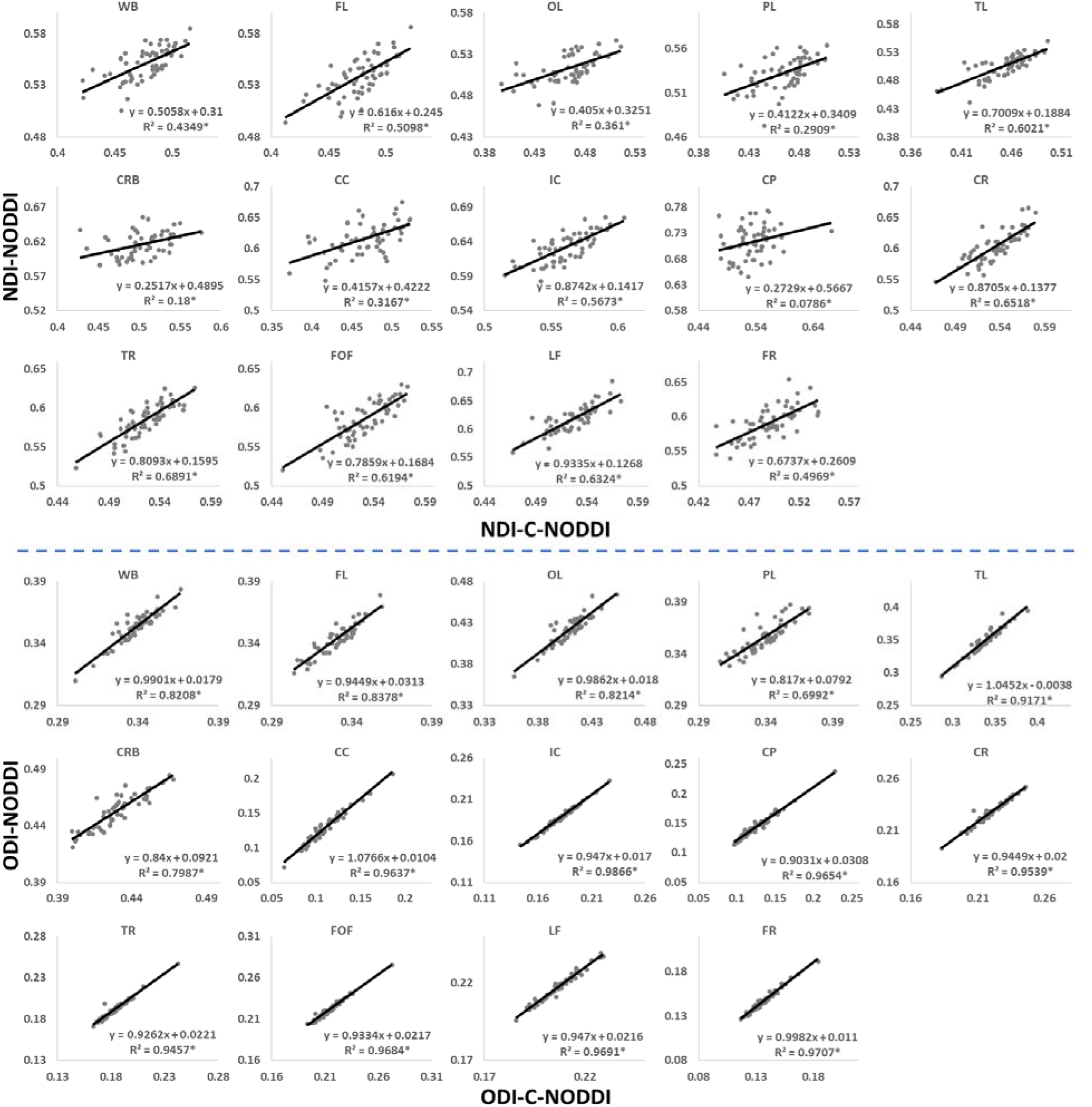
Pearson correlations between NDI, or ODI, derived values using C-NODDI and NODDI in the 14 WM ROIs studied. The coefficient of determination is provided with * indicating significant correlation. While derived ODI values using NODDI or C-NODDI were virtually identical exhibiting very high correlation coefficients, the NDI values derived using these two approaches were substantially different with NODDI exhibiting abnormally high NDI values in several WM regions. WB: whole brain, FL: frontal lobes, PL: parietal lobes, TL: temporal lobes, OL: occipital lobes, CRB: cerebellum, CC: corpus callosum, IC: internal capsules, CP: cerebral peduncle, CR: corona radiata, TR: thalamic radiation, FOF: fronto-occipital fasciculus, LF: longitudinal fasciculus, FR: forceps.

Figures 4a and 4b show, respectively, quantitative results for the NDI and ODI values calculated using NODDI or C-NODDI from all participants as a function of age for the indicated 14 WM regions. These results show, in agreement with Figure 3, increasing NDI until middle age followed by decreases afterward in most examined ROIs, with the best-fit curves displaying regional variation. Remarkably, the quadratic effect of age on NDI derived using C-NODDI was readily observable in almost all ROIs, as compared to the NDI results derived using NODDI which was limited to a few. Furthermore, in agreement with Figure 3, ODI values derived using either NODDI approach exhibited similar regional trends that were, overall, constant with age with limited ROIs exhibiting either increasing or decreasing trends. In agreement with visual inspection, statistical analysis indicates that the quadratic effect of age, age^2^, on NDI derived using C-NODDI was significant (*p* <.05) or close to significance (*p* < .1) in several ROIs, while this effect on NDI derived using NODDI was limited to a few ROIs (Table 1). For both NODDI approaches, the quadratic effect of age on ODI was not significant. In contrast, the effect of age on NDI-C-NODDI was significant, or close to significance, in almost all ROIs, while the age effect on NDI-NODDI was observed in only three ROIs. For both NODDI approaches, the effect of age on ODI was only significant in very limited cerebral regions. In addition, the effect of sex on NDI or ODI was, overall, not significant. Finally, in all ROIs, the NDI-C-NODDI results exhibited peak values at younger ages, as compared to the NDI results derived using NODDI.

**Figure 4a.**
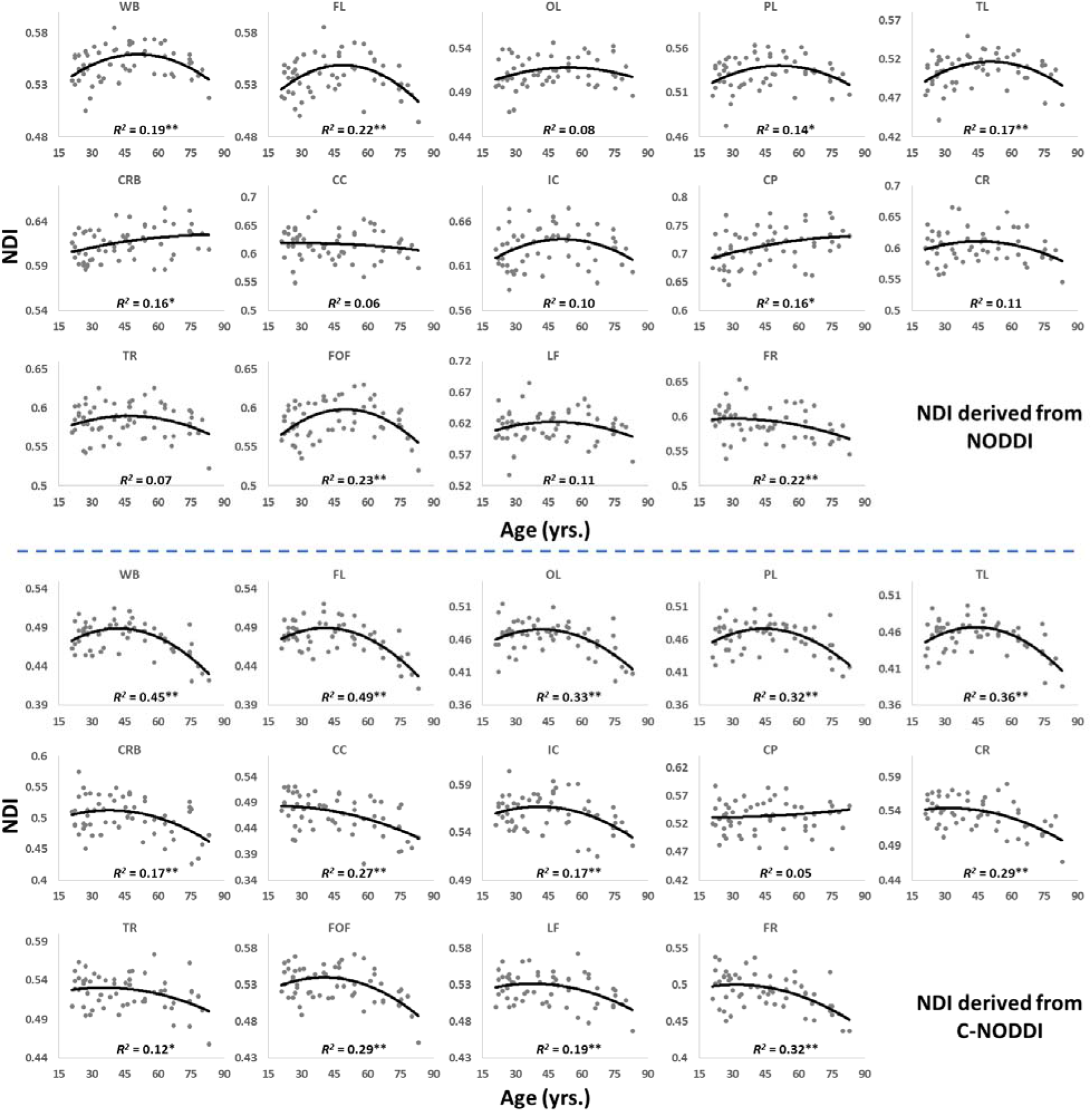
Regional NDI values derived using NODDI (top plots) or C-NODDI (bottom plots) as a function of age. Although most regions investigated exhibit inverted U-shaped trends of NDI with age, these quadratic trends were more noticeable and significant for the NDI-C-NODDI results (see Table 1). WB: whole brain, FL: frontal lobes, PL: parietal lobes, TL: temporal lobes, OL: occipital lobes, CRB: cerebellum, CC: corpus callosum, IC: internal capsules, CP: cerebral peduncle, CR: corona radiata, TR: thalamic radiation, FOF: fronto-occipital fasciculus, LF: longitudinal fasciculus, FR: forceps. * indicates p < 0.05 while ** indicates p < 0.01.

**Figure 4b.**
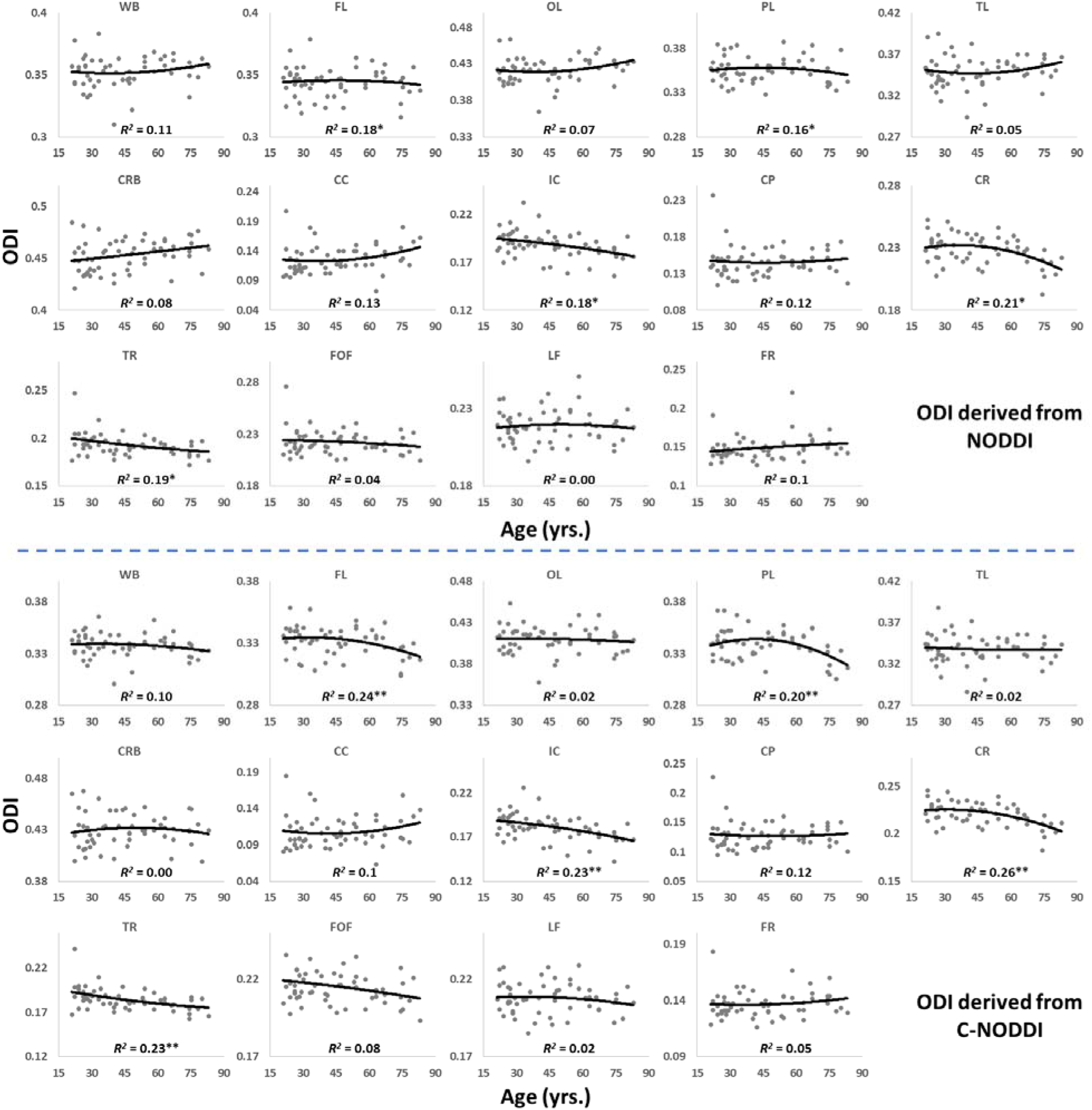
Regional ODI values derived using NODDI (top plots) or C-NODDI (bottom plots) as a function of age. Results were very similar for both approaches. WB: whole brain, FL: frontal lobes, PL: parietal lobes, TL: temporal lobes, OL: occipital lobes, CRB: cerebellum, CC: corpus callosum, IC: internal capsules, CP: cerebral peduncle, CR: corona radiata, TR: thalamic radiation, FOF: fronto-occipital fasciculus, LF: longitudinal fasciculus, FR: forceps.

**Table 1.**
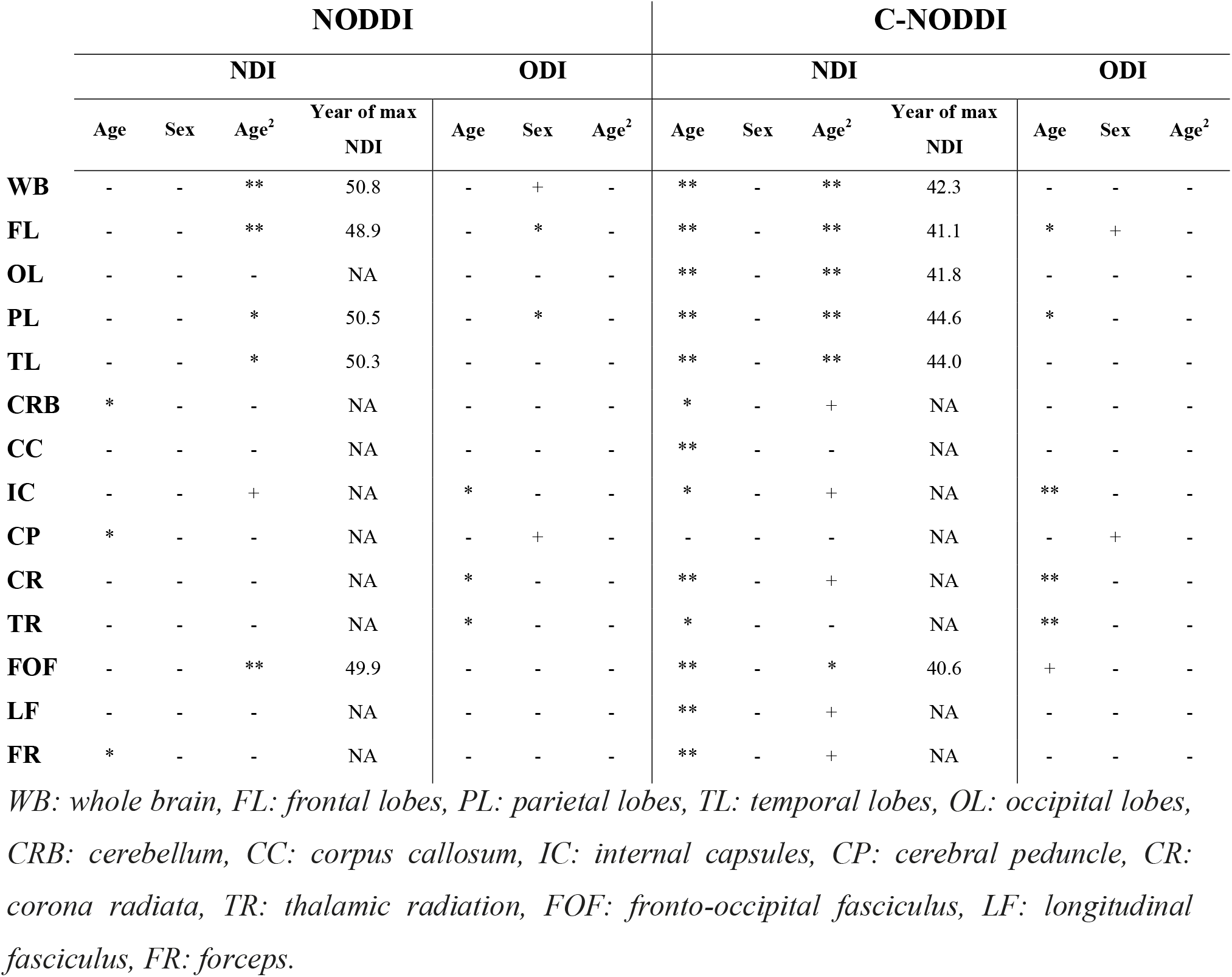
Significance of each coefficient incorporated into the linear regression analysis of NDI or ODI derived using NODDI or C-NODDI, and the year of apparent maximum NDI in each ROI. NA indicates “not applicable” for a nonsignificant age^2^ term. * Indicates p < .05, ** indicates p < .01, + indicates p < .1, and – indicates non-significant effects. All p-values presented are obtained after FDR correction.

|  | NODDI |  |  |  |  |  |  | C-NODDI |  |  |  |  |  |  |
| --- | --- | --- | --- | --- | --- | --- | --- | --- | --- | --- | --- | --- | --- | --- |
|  | NDI |  |  |  | ODI |  |  | NDI |  |  |  | ODI |  |  |
|  | Age | Sex | Age<sup>2</sup> | Year of max NDI | Age | Sex | Age<sup>2</sup> | Age | Sex | Age<sup>2</sup> | Year of max NDI | Age | Sex | Age<sup>2</sup> |
| **WB** | - | - | ** | 50.8 | - | + | - | ** | - | ** | 42.3 | - | - | - |
| **FL** | - | - | ** | 48.9 | - | * | - | ** | - | ** | 41.1 | * | + | - |
| **OL** | - | - | - | NA | - | - | - | ** | - | ** | 41.8 | - | - | - |
| **PL** | - | - | * | 50.5 | - | * | - | ** | - | ** | 44.6 | * | - | - |
| **TL** | - | - | * | 50.3 | - | - | - | ** | - | ** | 44.0 | - | - | - |
| **CRB** | * | - | - | NA | - | - | - | * | - | + | NA | - | - | - |
| **CC** | - | - | - | NA | - | - | - | ** | - | - | NA | - | - | - |
| **IC** | - | - | + | NA | * | - | - | * | - | + | NA | ** | - | - |
| **CP** | * | - | - | NA | - | + | - | - | - | - | NA | - | + | - |
| **CR** | - | - | - | NA | * | - | - | ** | - | + | NA | ** | - | - |
| **TR** | - | - | - | NA | * | - | - | * | - | - | NA | ** | - | - |
| **FOF** | - | - | ** | 49.9 | - | - | - | ** | - | * | 40.6 | + | - | - |
| **LF** | - | - | - | NA | - | - | - | ** | - | + | NA | - | - | - |
| **FR** | * | - | - | NA | - | - | - | ** | - | + | NA | - | - | - |

*WB: whole brain, FL: frontal lobes, PL: parietal lobes, TL: temporal lobes, OL: occipital lobes, CRB: cerebellum, CC: corpus callosum, IC: internal capsules, CP: cerebral peduncle, CR: corona radiata, TR: thalamic radiation, FOF: fronto-occipital fasciculus, LF: longitudinal fasciculus, FR: forceps.*

Figures 5, 6 and 7 show, respectively, examples of derived NDI and ODI maps, derived difference maps, mean parameter values calculated over the whole WM region, and Ratio-BIC maps, using NODDI or C-NODDI at several TEs using DWI data acquired from the brains of three participants. Visual inspection and quantitative analysis indicate that derived NDI estimates using NODDI are TE-dependent exhibiting increased values with TE increases. In contrast, the NDI maps and mean values calculated using C-NODDI are essentially similar with the difference maps exhibiting values close to zero in most white matter regions. However, although the large majority of the white matter regions exhibited relatively lower dependence on TE, some regions showed substantially increased values with TE. Further, it is readily seen that the ODI maps calculated using NODDI or C-NODDI are similar and with mean values virtually constant or exhibiting slightly decreasing trends as a function of TE. Moreover, we note that derived NDI and ODI maps exhibit some regional differences across TEs; this is due to differences in signal-to-noise ratio due to the large differences in TEs leading to signal drops at higher TEs, as well as to some persisting image misregistration. Further, the Ratio-BIC maps exhibited values that are mostly very close to or marginally lower than 1 in a large number of white matter regions suggesting that the parsimonious model, C-NODDI, would be preferred as compared to NODDI in these brain regions. However, we note that several other regions exhibited Ratio-BIC values that are higher than one implying that NODDI provided a better fit in these voxels. Finally, our Pearson correlation between NDI and NfL indicates stronger correlations between NDI-C-NODDI vs. NfL as compared to NDI-NODDI vs. NfL, with lower NDI values corresponding to higher NfL concentration levels (Fig. 8).

**Figure 5a.**
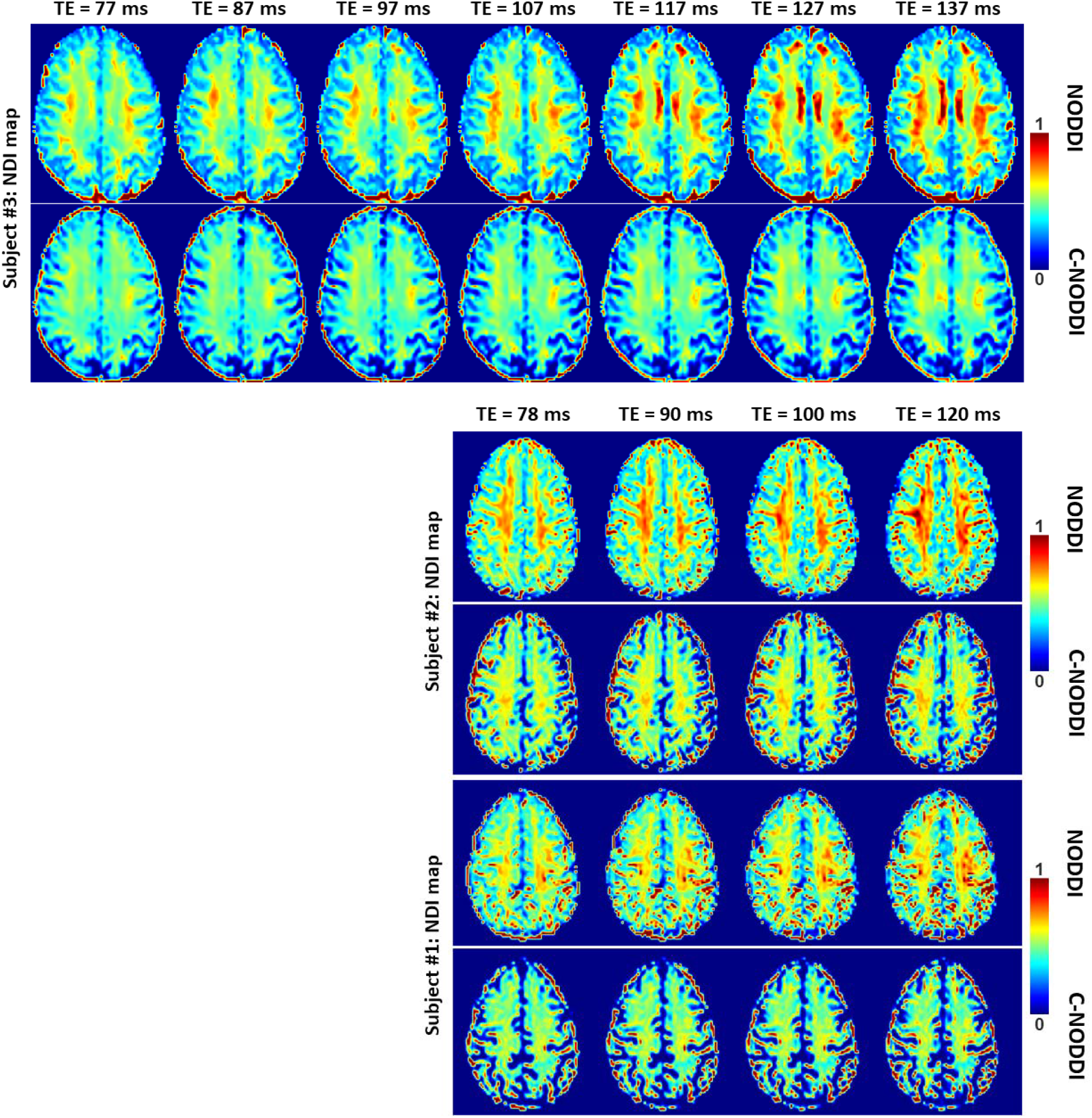
NDI maps derived using NODDI or C-NODDI from DWI data acquired at different TEs from the brains of three different subjects. Results are shown for a representative slice for each of the three participants. Visual inspection indicates that the regional NDI values derived using NODDI increase with TE, while remaining relatively constant using C-NODDI.

**Figure 5b.**
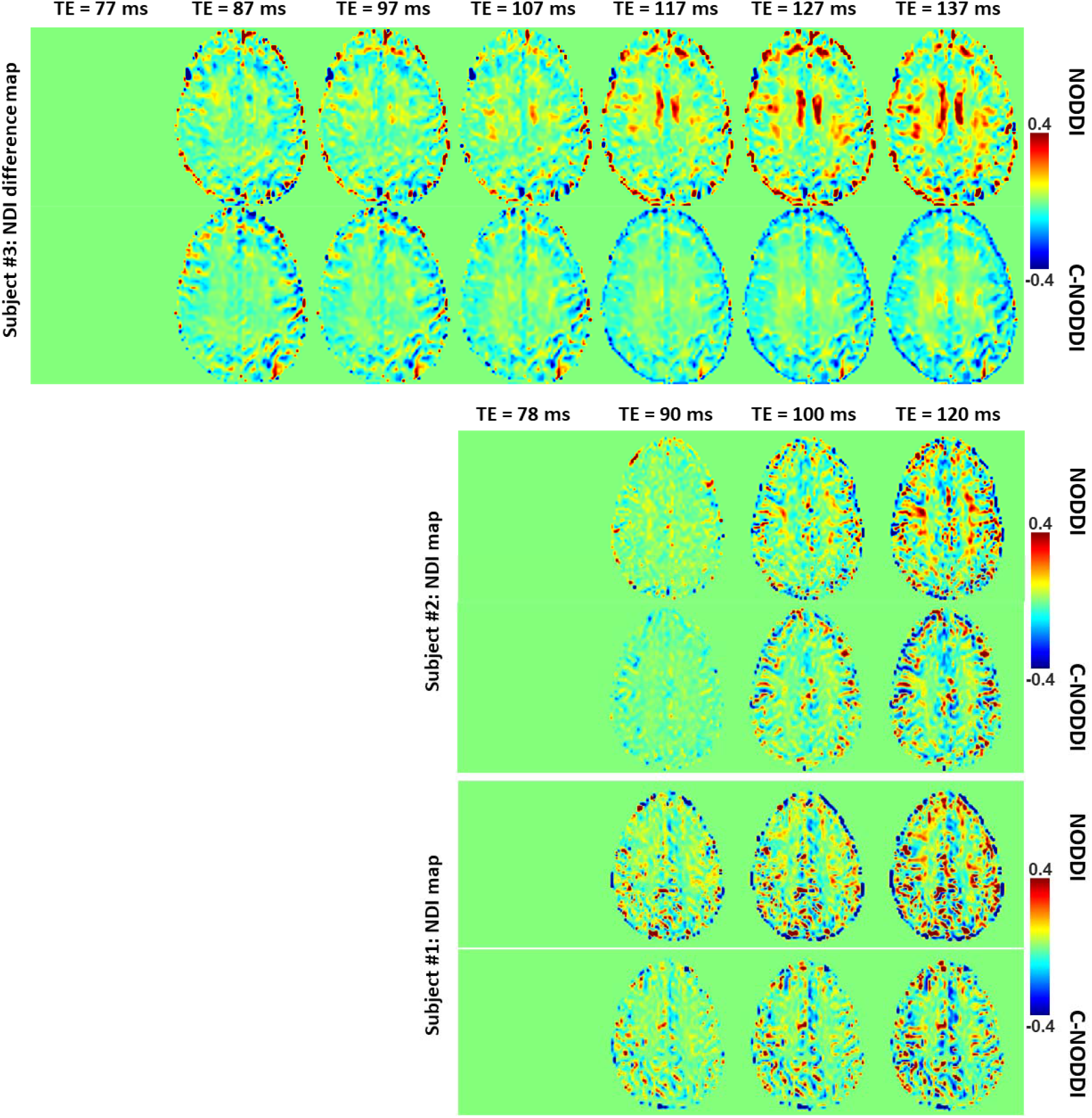
NDI difference maps calculated at different TEs from the brains of three different subjects. The NDI map derived at the minimal TE was used as the reference map. Results are shown for a representative slice for each of the three participants. It is readily seen that derived difference values were near zero in most white matter regions using C-NODDI while exhibiting large positive values in several white matter regions using NODDI.

**Figure 5c.**
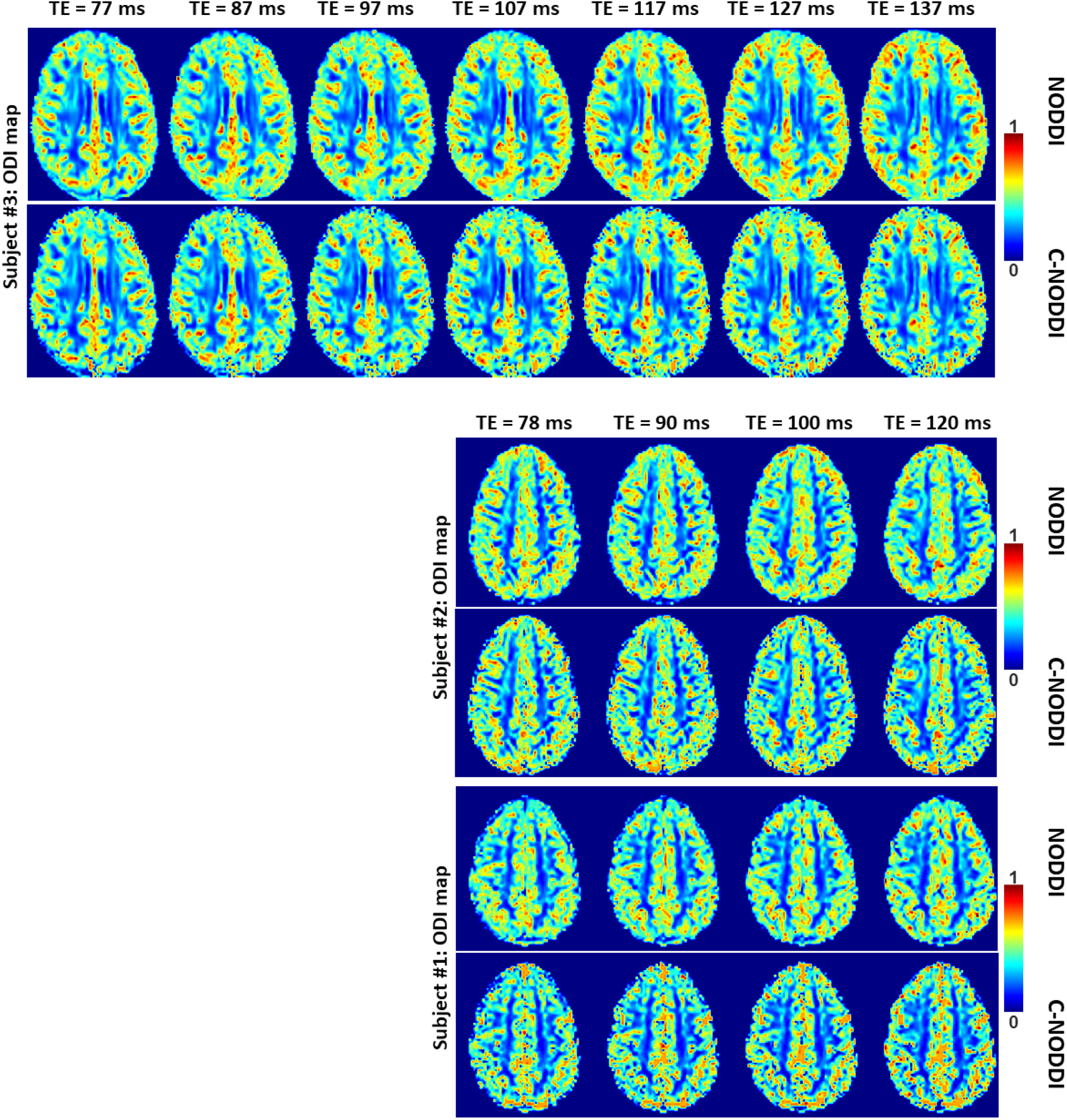
ODI maps derived using NODDI or C-NODDI from DWI data acquired at different TEs from the brains of three different subjects. Results are shown for a representative slice for each of the three participants. Visual inspection indicates that the regional derived ODI values using either approach are virtually identical as a function of TE.

**Figure 5d.**
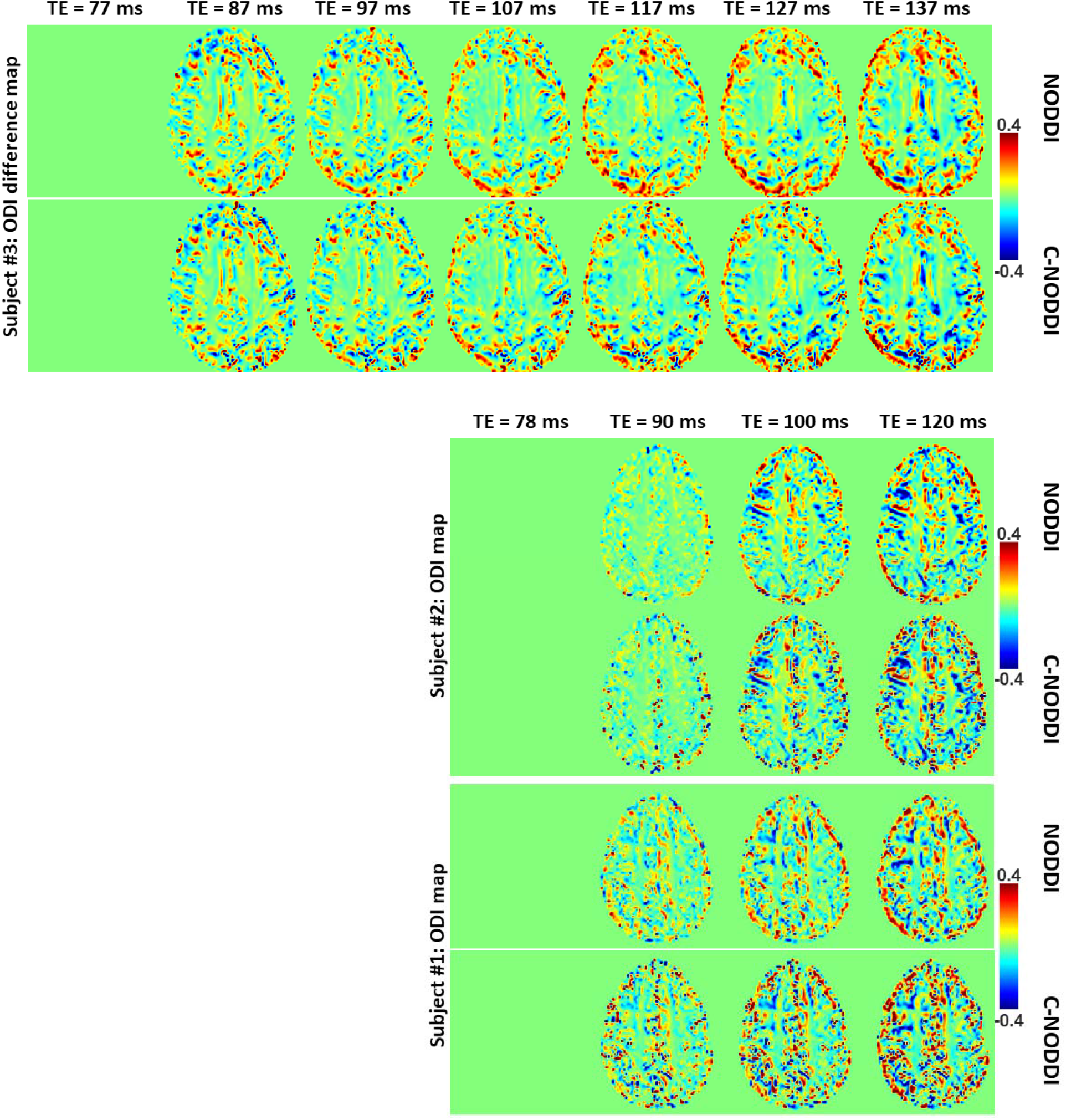
ODI difference maps calculated at different TEs from the brains of three different subjects. The ODI map derived at the minimal TE was used as the reference map. Results are shown for a representative slice for each of the three participants. It is readily seen that derived difference values were near zero in most white matter regions using either approach.

**Figure 6.**
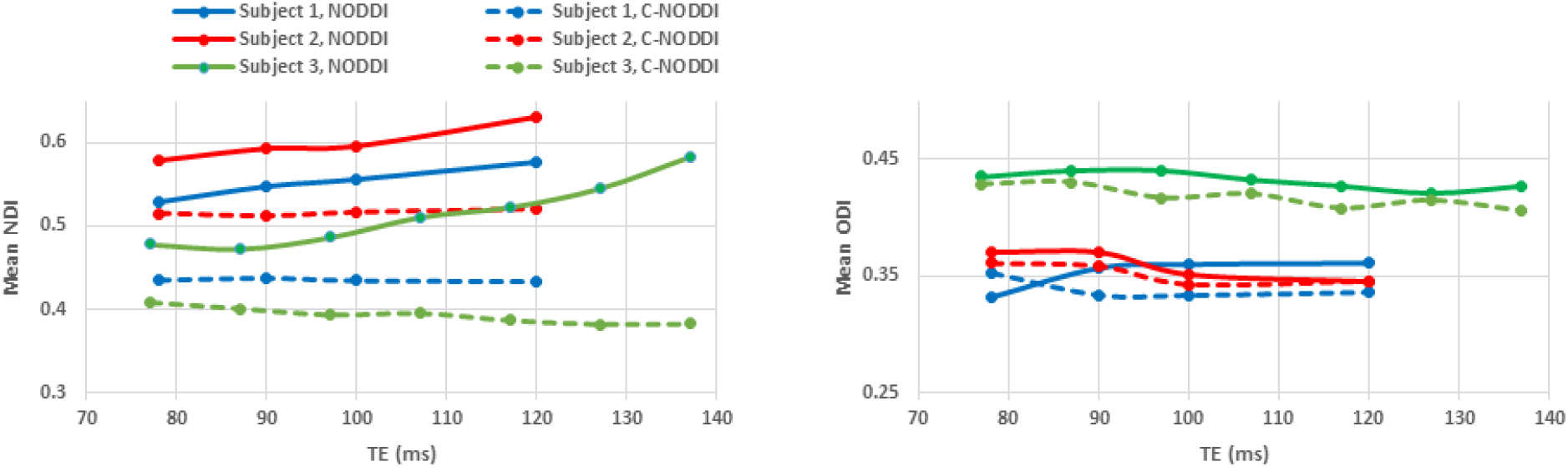
Mean NDI values (A) and mean ODI values (B), calculated from a large WM region encompassing the cerebral lobes, derived using NODDI (red) or C-NODDI (blue) as a function of TE. It is readily seen from this quantitative analysis that the NDI values derived using NODDI increase with TE, while derived ODI values using either approach are relatively similar between both approaches with, overall, constant or decreasing trends as a function of TE, in agreement with the visual inspection (Fig. 5).

**Figure 7.**
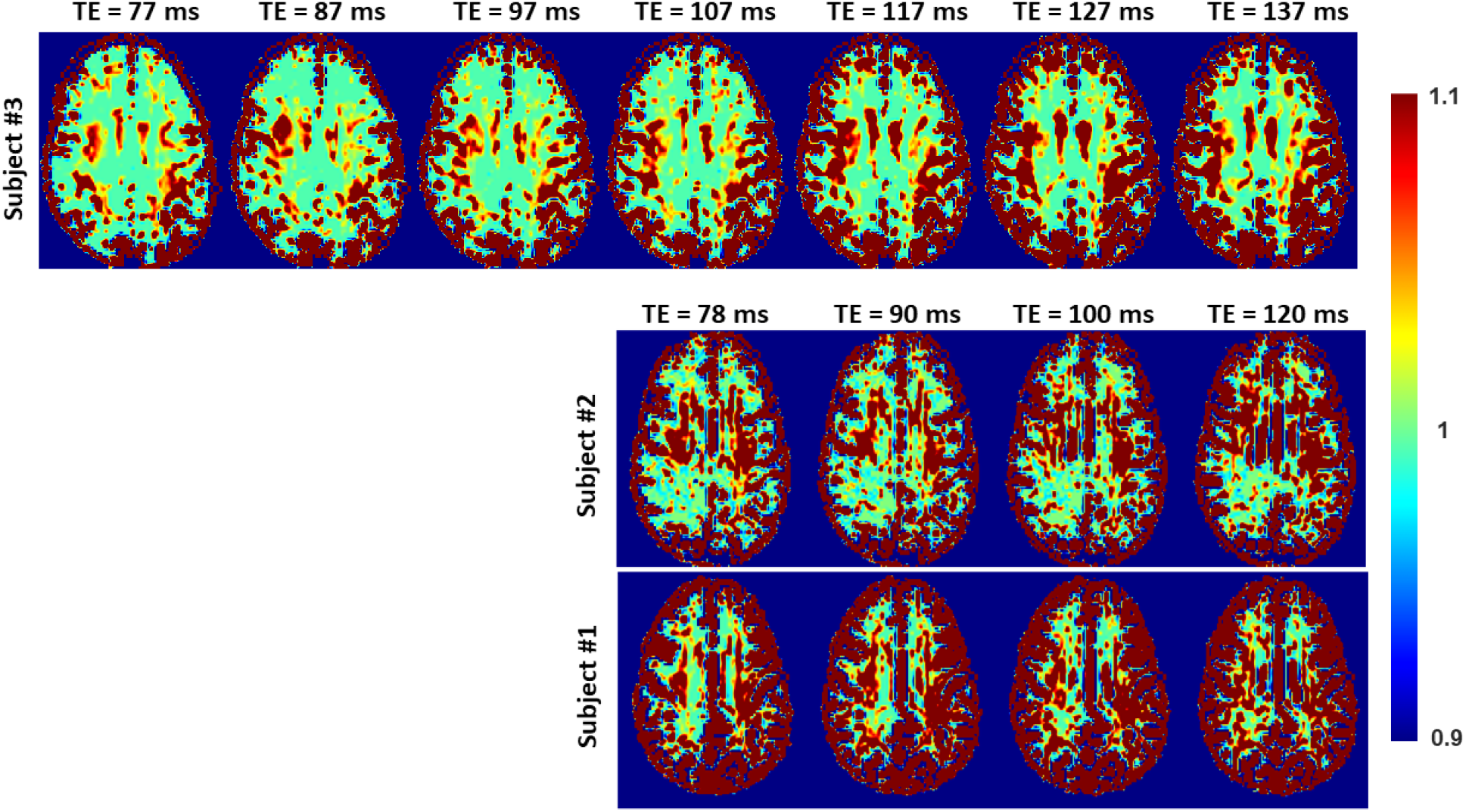
Representative examples of the Ratio-BIC maps, defined as the ration between BIC-C-NODDI and BIC-NODDI, from the brain of the three subjects.

**Figure 8.**
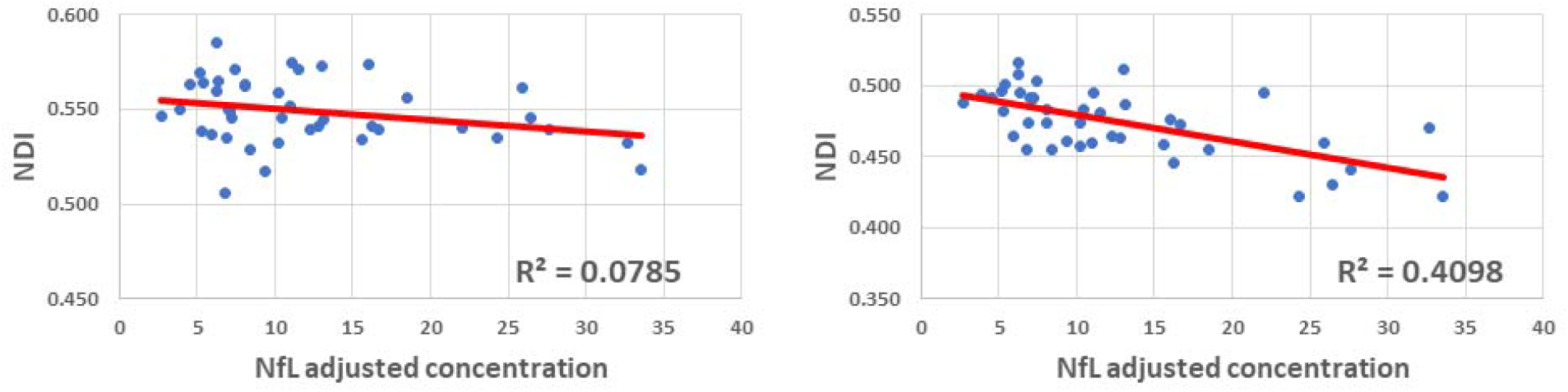
Correlation plots between neurodegeneration biomarker, NfL, and whole brain white matter brain NDI values derived using NODDI (left plot) or C-NODDI (right plot). Line of fit (in red) and R^2^ values were displayed.

Figure 9 shows NDI maps derived using MTE-NODDI from the brains of three different subjects. Visual inspection indicates that, while the regional NDI values derived using MTE-NODDI were, overall, higher and greatly dispersed as compared to those derived using either NODDI or C-NODDI for both Subject #1 and Subject #2 (Fig. 5a), the derived regional NDI values using MTE-NODDI were very similar to those obtained using C-NODDI for Subject #3 (Fig. 5a). The mean NDI values obtained in a large WM region encompassing the cerebral lobes using MTE-NODDI from Subject #3 was 0.41; this is virtually identical to the values derived using C-NODDI for different TEs (Fig. 6).

**Figure 9.**
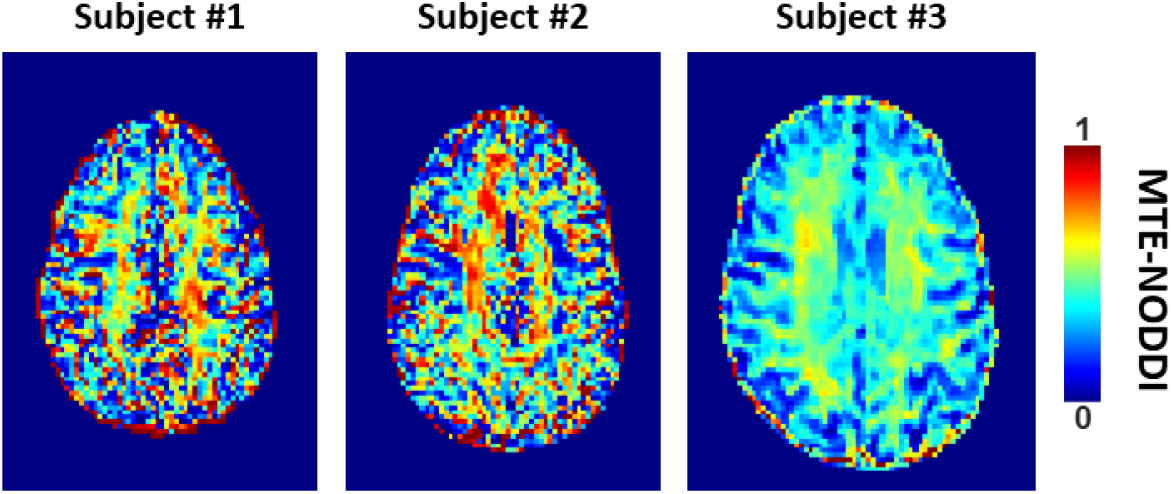
NDI maps derived using MTE-NODDI from DWI data acquired at different TEs from the brains of three different subjects. Results are shown for a representative slice for each of the three participants. Visual inspection indicates that while the regional NDI values derived using MTE-NODDI exhibit overall large values for both Subject #1 and Subject #2 as compared to those derived using either NODDI or C-NODDI, derived regional NDI values using MTE-NODDI were very similar to those obtained using C-NODDI for Subject #3.

## DISCUSSION & CONCLUSIONS

In this work, we introduced a modification of the NODDI signal model, C-NODDI, to overcome several documented problems with the original NODDI approach. Specifically, this modification addresses the overestimations of the CSF and NDI fractions in WM, as well as the dependency of derived NDI values on TE. Our results show that C-NODDI provides lower NDI estimates as compared to NODDI and confirm that NODDI provides high NDI values in several WM structures with values over 70% at several cerebral structures in agreement with previous reports (41, 42). NDI values derived from C-NODDI were regionally 20% to 40% lower than those derived using NODDI; this agrees with Bouyagoub and colleagues’ results (42). However, unlike their approach which requires a substantial extension of the total acquisition time, our approach does not require additional acquisition time. Indeed, C-NODDI is based on prior estimation of the signal fraction of the CSF. In this proof-of-concept work, we showed that this fraction can be estimated using the *T_2_*-weigthed image acquired at *b*-value of 0 s/mm^2^ and the hidden Markov random field model and the expectation-maximization algorithm; this allows reduction of the fitting parameter space of the NODDI model (63). Indeed, the NODDI signal model incorporates several parameters to be estimated. Parameter estimation in high dimensional problems, that is, those for which the number of parameters to be estimated is large, is complicated by the presence of local minima and saddle points (53). This problem becomes more acute with the flatness of the least-squares residual surfaces seen with increasing model complexity exhibiting higher sloppiness, that is, different parameter combinations leading to virtually equivalent signal behavior (50–52). However, we note that both NODDI and C-NODDI signal models are just approximations of the underlying biology so that further histological validations are required to assess their reliability and accuracy. We note that the CSF fraction can be estimated using different approaches, including the free-water DTI approaches from the same NODDI dataset, or using FAST from various other structural contrast-weighted images that are routinely acquired in clinical studies (64, 65). A thorough comparison of all these techniques is still needed, especially in the context of their applications to neurodegeneration.

Furthermore, using data acquired from three participants at different TEs, we confirmed Gong and colleagues’ observation that the NDI estimates are dependent on TE, with NDI values increasing with TE (41). To address this concern, Gong and colleagues suggested a multi-TE NODDI approach based on acquiring several NODDI images at different TEs to account for the *T_2_*-weigthing of each of the three compartments incorporated in the NODDI signal model (41).

Indeed, in MTE-NODDI, T*_2,in_*, T*_2,ex_* and T*_2,iso_* are considered to have different values, including in the WM regions; this is consistent with previous observations (66). However, this assumption disagrees with extensive relaxometry-based evidence that the transverse relaxation times of the intracellular and extracellular waters have close values, especially in healthy WM (45, 46, 67-75). Our results presented here somehow support this observation. Indeed, derived NDI values using C-NODDI exhibited a lower dependence on TE in several WM structures. We believe that the artificial overestimation of the CSF fraction using NODDI is a leading factor of the strong underlying TE-dependency observed previously, particularly in WM. Further, derived CSF water fraction using MTE-NODDI remains high, exhibiting values near 20% in several deep WM regions. This is physiologically not plausible and clearly indicates that the MTE-NODDI method could not resolve this outstanding issue in the NODDI signal modeling. In fact, this limitation of the MTE-NODDI method motivated our proposed C-NODDI approach, which overcomes this issue. However, we note that some white matter regions showed dependence on TE, exhibiting relatively higher values with increasing TE. However, further examination of the effect of T_2_s on derived diffusion parameters is required to shed light on the mechanisms underlying this discrepancy. Interestingly, despite these differences between approaches, MTE-NODDI and C-NODDI exhibited similar NDI values, especially when the number of DWI images was obtained with a larger number of TEs and *b*-values; this agrees with Gong and colleagues’ observation of lower NDI values using MTE-NODDI as compared to those derived using NODDI (41). However, when the number and range of TEs were limited, the MTE-NODDI exhibited very different and highly dispersed NDI values as compared to those derived using either NODDI or C-NODDI. This observation motivates a comprehensive comparison between these approaches at different experimental designs, including in terms of different ranges and numbers of TEs and *b*-values; this work is underway. Finally, in perfect agreement with Bouyagoub’s and Gong’s results (41, 42), the ODI values derived using NODDI or C-NODDI were virtually identical, as expected. Indeed, the calculation of ODI does not incorporate signal fractions and thus remains insensitive to differences in the CSF fraction.

We investigated the association between NDI or ODI, derived using NODDI or C-NODDI, and age in several cerebral WM structures in a healthy adult population spanning a wide age range. Our results revealed widespread WM microstructural differences as a function of age, as well as regional variations between the NDI or ODI measures and age. Specifically, NDI exhibited quadratic, inverted U-shaped, regional trends with age; this agrees with our and Beck and colleagues’ recent results (6, 7). We attribute this finding to a continuous maturation of axons with increases in axonal density until middle age, followed by a phase of neurodegeneration and consequence axonal loss at older ages. Interestingly, the quadratic effect of age, age^2^, on NDI was significant (*p* < .05) or close to significance (*p* < .1) in most regions investigated using C-NODDI, while this inverted U-shaped association was limited to only a few cerebral regions using the original NODDI approach. Although this observation indicates the potential higher sensitivity of C-NODDI to capturing differences in neurite density with age, further comparison between NODDI and C-NODDI in larger cohorts is still required to confirm or infirm this finding. We note that other investigations have shown either higher or lower NDI values with age in WM (38–40). This discrepancy is likely due to differences in the characteristics of the study cohorts as well the instability of NODDI. Indeed, Billiet’s and Chang’s studies, although incorporating wide age ranges, possess only a limited number of subjects over 60 years old, while Merluzzi’s cohort included only subjects over 45 years old. These limitations may have precluded the detection of the quadratic association between NDI and age, as Beck and we have observed (6, 7). This issue may have been exacerbated by differences in the experimental implementation of NODDI given its high sensitivity to TE (41). Further, ODI exhibited non-consistent regional trends with age, with most WM regions exhibiting constant trends while other, but limited, regions exhibited either increasing or decreasing trends. The literature regarding differences in ODI with age remains sparse, necessitating further detailed investigations (6, 38–40). Finally, our results revealed that elevated concentration levels of NfL were associated with lower NDI values suggesting a role of axonal degeneration in neuroinflammation. This association was stronger between NDI derived using C-NODDI and NfL, suggesting that NDI-C-NODDI could represent a reliable imaging biomarker of axonal integrity. Further studies in larger cohorts and using NfL concentrations derived from CSF are still required.

In all ROIs, NDI calculated using C-NODDI exhibited peak values at much younger ages compared to NDI calculated using NODDI, with differences ranging from ∼6 to ∼10 years. This indicates that axonal maturation continues until the early fifth decade of age. This finding agrees with results derived using sensitive MRI measures of axonal density, including relaxation times and DTI indices, all indicating that WM tissue maturation continues until the late fourth decade to the early fifth decade of age (14, 15, 17, 76). Interestingly, recent studies have shown that myelination continues until the early fifth decade of age as well (16, 77, 78). It has been shown that axonal activity is an important contributing factor in myelin modulation (79, 80). Moreover, aside from acting as an electric insulator, oligodendrocytes, the cells that produce myelin, provide substantial metabolic support to axons (81). Given this physiological coupling between axons and myelination, it is plausible that axonal maturation and myelin maturation follow similar patterns and peak at similar ages. However, further investigations on larger cohorts, in addition to longitudinal studies, are required to derive definite conclusions regarding whether the maturation and myelination pattern of axons are mechanistically associated or represent two independent neurological processes. These studies are crucial for the development of specific interventions supporting myelin maintenance, axonal regeneration, or both simultaneously.

Our work has limitations. Although our cohort spans a wide age range, it does not include participants under the age of 20 due to the inclusion and exclusion criteria of the BLSA and GESTALT studies. Including younger participants may influence the shape of the NDI or ODI age-related trends and the assessment of their respective maxima with respect to age (82). With our current dataset being cross-sectional, our results could be further validated through longitudinal studies; such work is underway. In addition, the NODDI and C-NODDI implementations are based on several assumptions and fixed values for certain diffusivity parameters to improve the stability of the signal model. Further analyses are needed to investigate the effects of these fixed parameters on the NODDI and C-NODDI derived parameters. Moreover, our acquisition protocol included only one image at *b*-value of 0 s/mm^2^.

However, it has been shown that several *b_0_* images are critical for an accurate determination of diffusion parameters (83). Finally, our method was applied to cognitively unimpaired individuals with further investigations required in the context of neurodegeneration and cognitive impairment.

## Data Availability

All data produced in the present study are available upon reasonable request to the authors

